# Predictors of Breast Self-Examination Practice Among Women Aged 20 and Above In Sidama Region, Southern Ethiopia. Health Belief Model Approach

**DOI:** 10.1101/2023.12.27.23300590

**Authors:** Belayneh Bekele, Ermias Yunkura, Netsanet Bogale, Lelisa Gemechu, Achamyelesh Gebretsadik, Dereje Geleta

## Abstract

**Introduction:** Breast self-examination is a straightforward, affordable, and uncomplicated approach to identify any alteration in the breast. When conducted correctly, it aids in the early detection of breast cancer and diminishes its impact on health and mortality. It is advised that women aged 20 and above perform self-examinations monthly, consistently. However, the extent of breast self-examination practices in Ethiopia has not been thoroughly documented.

**Objective:** The aim of the study is to identify predictors of breast self-examination practices and associated factors among women aged 20 and above in Sidama Region, South Ethiopia.

**Methods:** A Cross-sectional study was conducted on 1000 women aged 20-70 years. The study employed a multistage sampling method. Data collection was performed using a structured and pretested interviewer administered questionnaire. The collected data were entered into Epi-data version 3.1 and analyzed by SPSS version 20. Descriptive statistics were used to describe the study population in relation to relevant variables. Bivariate and multivariable logistic regression analyses were conducted to assess the impact of independent variables on the dependent variable. Variables with p-value <0.25 were considered as candidates for multivariable logistic regression. Statistical significance was declared at P < 0.05 and 95% CI.

**Result:** The prevalence of breast self-examination was found to be 5.2%, with only 46.2% of participants performing it regularly. Factors significantly associated with the practice of BSE among women included were Good knowledge about breast cancer and breast self-examination (Adjusted odds ratio=4.5, 95%CI: 1.4, 14.7), high perceived susceptibility (Adjusted odds Ratio =2.7, 95%CI: 1.4, 5.2), high perceived barrier (Adjusted odds ratio=0.30, 95%CI: 0.11, 0.8) and high perceived self-efficacy (Adjusted odds ratio=3.9, 95%CI: 2.0, 7.7).

**Conclusion:** The study revealed a notably low prevalence of BSE practice among the study population compared to previous study conducted in Ethiopia. Factors such as knowledge about breast cancer and breast self-examination, high perceived susceptibility, and high perceived self-efficacy were identified as significant predictors of BSE practice. To address this issue, collaboration and efforts from various stakeholders including Zonal Health department, District health offices, Health workers, and other relevant parties are needed. It is crucial to focus on raising awareness and improving the perception of women regarding BSE to enhance the currently inadequate practice.

## Background

Breast cancer is a cancer type which starts in breast cell(1). It is a significant health concern in women all over the globe; and the second most common cancer in both the industrialized and developing world overall (2).

It is the most commonly diagnosed disease in women and the leading cause of cancer death globally, the incidence of breast cancer is increasing in the developing world due to the adoption of western lifestyles, increased life expectancy and increasing urbanization(3).

Globally with an estimated 2.1 million new cases and 627,000 deaths in 2018-that is, around 15% of all cancer death among women(4). It also the primary cause of death in the less developed countries of the world, and an emerging public health problem in Sub-Saharan Africa(5,6).

In Ethiopia, breast cancer is the second common cancer next to cervical cancer. Around 10,000 breast cancer cases diagnosed in late stage annually, although a high number of cases live in the society who don’t seek health service in the early stage(7).

In the limited resource setting, late diagnosis of breast cancer is common. More than 70% of patients visit a health facilities with an advanced stage of the disease. The delayed presentation has been associated with a low level of community and health professionals’ awareness of breast cancer, poor distribution of health infrastructure and inadequate supply, incomplete vital registration, low educational empowerment of women, delayed health-seeking behaviour due to strong traditional beliefs, and absence of detection programs. In the absence of systematic screening programs, in low resource countries (LRC), most breast cancers present with self-discovered “painless breast lump” (3,8)

To reduce the morbidity and mortality of breast cancer, early detection and effective treatment play an important role(9). Breast self-examination (BSE) is one of the most effective and feasible techniques for early detection of breast cancer since every woman aged 20 and above years has to be done for 20 minutes every month and is recommended primarily for a resource-limited country(1). However, in developing countries including Ethiopia, BSE practice remains low (10–12).

A study was conducted regarding BSE in Saudi women showed that 17% and 21.1% practice BSE (13,14). A study conducted in Nigeria showed that 18.1% practice BSE(15). A Study in Cameroon, only 3% regular performs BSE(16). Similarly, a study was conducted in Ethiopia, only 12% and 14.4% perform BSE(10,11). The main reason women do not perform BSE, lack of awareness, do not know how to perform, poor information accesses, low perceived susceptibility, low perceived severity, and low perceived benefit, do not know the importance and self-efficacy the most prominent factors (10–12,17).

However, the Ethiopian government currently develops a cancer prevention and controlling strategies. It focuses in particularly to prevent and control cancer by strengthening health promotion and education activities to prevent the individual themselves from modifiable risk factors and improvement of early detection of cancer by introducing or expanding the available screening program intervention and capacitate the existing health systems, but it is not applicable in the grass-root level still more priority given to communicable disease(18,19). Studies done on the issue in Ethiopia have been very limited. Moreover, the extent of BSE practice prevalent and recent data is scarce in South Ethiopia, particularly in the study setting, and this study aims to determine the level of BSE practice and its associated factors in Sidama Zone, South Ethiopia.

## Methods and materials

### Study area

The study was conducted in three randomly selected districts of Sidama Region.. According to 2007, Ethiopia central statistics agency (CSA) census, the estimated population estimate for the year 2018/2019 is approximately 3,893,816 to 1,939,120 are males and 1,954,696 of them are females. Among the total population, approximately 694,234 women are aged 20 years and above.

### Study design

The community-based cross-sectional study design was conducted

### Study unit

The study participants were randomly selected women aged 2o years and above who resided in the households of the selected kebeles. Data for the study were collected directly from these women.

### Measurements

#### BSE practice

A woman who performs BSE is regular at least once in a month. In this study, the practice of BSE is considered to perform breast examination on their breast at least ones in a lifetime(20).

#### Knowledge

There were 14 multiple choice questioners that carried out a total of 19 correct responses that measure knowledge. Each correct response was given a score 1 and a wrong response score 0.

### According to Bloom’s classification, cut off points for knowledge score(20)

1. Good knowledge a score of 80-100%
2. Satisfactory knowledge with a score of 60-70%
3. Poor knowledge, and a score of less than 60% of the correct response.

#### Perception on BSE

The beliefs and feeling of the respondent (who should BSE for breast cancer, intention to use BSE in the future). The scoring system used concerning respondents’ responses was assessed as follows: Strongly agree score 5, agree 4, neutral 3, disagree 2, strongly disagree 1. All scales are positively related to screening behaviour, except for barriers which are negatively associated.

### Sample size determination and sampling approach

The sample size for this study was determined using the single population proportion formula, considering a 12% proportion of breast self-examination (BSE) practice based on previous research (11). With a 3% margin of error, a 95% confidence level, a 10% nonresponsive rate, and a design effect of 2, the final sample size was calculated to be 992. However, this study was part of a larger thematic research project conducted by Hawassa University School of Public Health titled “Cancer in Southern Ethiopia: Epidemiological, psychosocial, and Economic Burden,” the sample size of 1000 from the thematic study was adopted for this research as well.

The selection of study areas involved a random selection of three woredas through a lottery method. Subsequently, 13 kebeles were chosen from each of the three woredas using the same procedure. The calculated sample size was then proportionally allocated to each selected woreda based on the number of women aged 20 years and above. A similar process was followed to determine the sample size for each selected kebele.

To identify households for the study, a list of households with women aged 20 years and above was obtained from the updated family folder in the health post. Systematic random sampling was employed, selecting every fourth household with eligible women for the interviews. In cases where multiple eligible women were present within a household, one woman was chosen randomly using a lottery method for the interview.

### Data collection

Data collection was conducted using an interviewer-administered structured questionnaire. The questionnaires were initially prepared in English and then translated into both Sidamu Afu and Amharic, the local languages spoken by the respondents in the study area. To ensure consistency of meaning, the translated Sidamu, afu, and Amharic versions of the questionnaire were translated back into English.

Prior to the main study, a pretest was carried out in selected kebeles that were not included in the main study. This was done to evaluate the feasibility and clarity of the questionnaire.

For data collection, a team of ten diploma nurses and two other health professionals with a degree were employed as data collectors and supervisors. These individuals were proficient in both Sidamu Afu and Amharic languages, enabling effective communication with the respondents.

### Data Analysis

The collected data were entered into EPI-data version 3.1 and subsequently exported to SPSS version 20 for statistical analysis. Descriptive statistical analysis was performed to calculate frequencies, percentages, and means for both independent and dependent variables.

In the bivariable analysis, variables with a significant association (p-value < 0.25) were selected and included in the multivariable analysis to identify independent predictors of BSE. The goodness of fit of the model was assessed using the Hosmer-Lemeshow statistic, with a not significant p-value of 0.82, and the omnibus test, which showed a significant p-value of less than 0.001. To account for possible confounding variables, multivariable logistic regression was employed. The COR and AOR were evaluated at 95% CI, and statistical significance was determined at a p-value < 0.05.

### Ethical considerations

The study received ethical approval from the IRB at Hawassa University, College of Medicine and Health Sciences. Prior to data collection, the purpose and objectives of the study were explained to the participants, and their informed consent was obtained. Participants were assured of their right to withdraw from the study at any time without facing any negative consequences. To protect participant confidentiality, coding was used to exclude personal identification information and names from the study data throughout the entire research process.

## Result

### Socio-demographic characteristics of respondents

A total of 1000 women participated in this study, resulting in a response rate of 100%. The mean age of the participants was 30.94 with [SD #x00B1; 6.63]. In terms of marital status, the majority of participants, 930 (93%), were married. Protestantism was the dominant religion among the participants, accounting for 825 (82.5%) individuals. Regarding educational background, 368 (36.8%) participants had not received any formal education, while the remaining had completed at least primary school or higher education.

In terms of occupation, the majority of participants, 811 (81.1%), were housewives. Other occupations included self-employment, which was reported by 99 (9.9%) participants, while 41 (4.1%) were students, and 4 (0.4%) were employed in private companies. (Table-1)

**Table 1:** Sociodemographic characteristics of women aged 20 and above years practice of breast self-examination in Sidama Region, South Ethiopia (N=1000)

| Variable | Frequency | Percent |
| --- | --- | --- |
| **Age** |  |  |
| 20-29 | 410 | 41 |
| 30-39 | 405 | 40.5 |
| 40 and above | 185 | 18.5 |
| **Marital status** |  |  |
| Single | 28 | 2.8 |
| Married | 930 | 93 |
| Divorced | 12 | 1.2 |
| Widowed | 22 | 2.2 |
| Separated | 8 | 0.8 |
| **Educational status of the women** |  |  |
| None formal education | 368 | 36.8 |
| primary education | 399 | 39.9 |
| Secondary education | 181 | 18.1 |
| Tertiary and above | 52 | 5.2 |
| **Educational status of the house head** |  |  |
| None formal education | 291 | 29.1 |
| Primary education | 369 | 36.9 |
| Secondary education | 249 | 24.9 |
| Tertiary and above | 91 | 9.1 |
| **Monthly income(in Eth birr)** |  |  |
| <445 | 134 | 13.4 |
| 445-2500 | 631 | 63.1 |
| 2501-3500 | 106 | 10.6 |
| 3501 and above | 129 | 12.9 |

### Family and personal history of breast cancer of the participants

Nearly all participants 993(99.3%), reported that they did not have a personal history of known breast cancer. Similarly, a majority of participants, 983(98.3%), stated that they did not have any family members who had been diagnosed with or passed away from breast cancer. However, a small proportion of the participants, 97 (9.7%), reported knowing someone suffering from breast cancer. (Table-2)

**Table 2:** Family and personal history of breast cancer among women aged 20 and above years practice of breast self-examination in Sidama Region, South Ethiopia (N=1000)

| Variable | Frequency | Per cent |
| --- | --- | --- |
| Personal history of breast cancer |  |  |
| No | 993 | 99.3 |
| Yes | 7 | 0.7 |
| Any family member died or sick of breast cancer |  |  |
| No | 983 | 98.3 |
| Yes | 17 | 1.7 |
| Who is affected or died by breast cancer(n=17) |  |  |
| Mother | 2 | 11.8 |
| Sister | 5 | 29.4 |
| Grandmother | 2 | 11.8 |
| Aunt | 8 | 47 |
| Know someone suffering from breast cancer |  |  |
| No | 903 | 90.3 |
| Yes | 97 | 9.7 |

### Knowledge of study participants on breast self-examination

The findings revealed that the majority of respondents, 686 (68.6%), had poor knowledge about BSE. Over half of the participants, 565 (56.5%), acknowledged that early detection of breast cancer increases the chance of survival. Furthermore, 478 (47.8%) participants were aware of the existence of breast cancer screening methods, while 447 (44.7%) knew about the specific types of screening methods available.

In terms of awareness about BSE, 418 (41.8%) participants reported having heard about it. Among the various sources of information, health professionals were the primary source, with 412 (41.2%) participants acquiring knowledge from them. This was followed by radio, which accounted for 129 (12.9%) participants, television with 95 (9.5%) participants, and health extension workers with 51 (5.1%) participants.(Figure-1)

**Figure 1:**
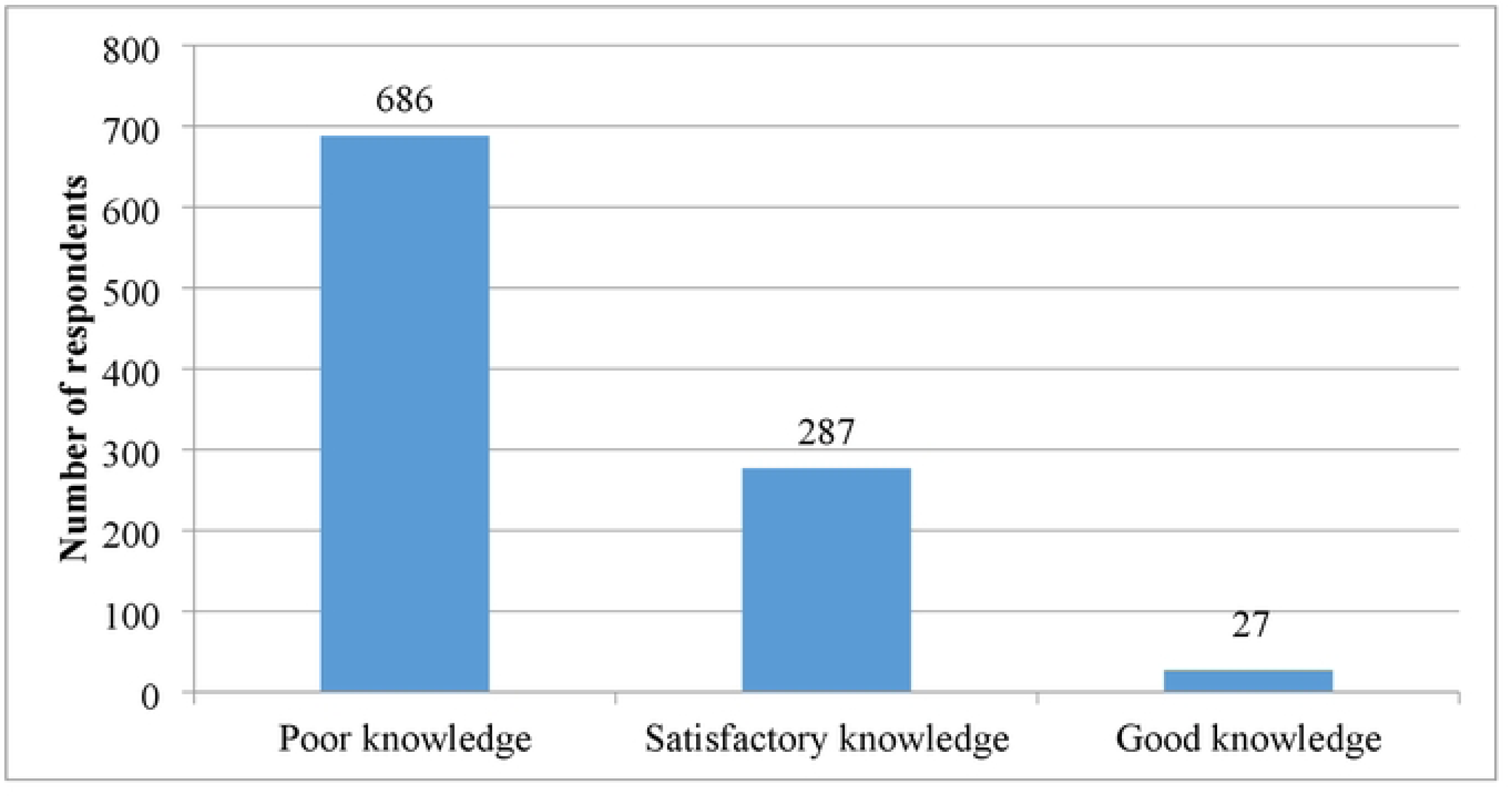
knowledge of breast self-examination among women aged 20 and above years

### Type of breast cancer screening methods

With regard to knowledge about breast cancer screening, 364(36.4%) of the respondents were familiar with BSE method. (Figure-2)

**Figure 2:**
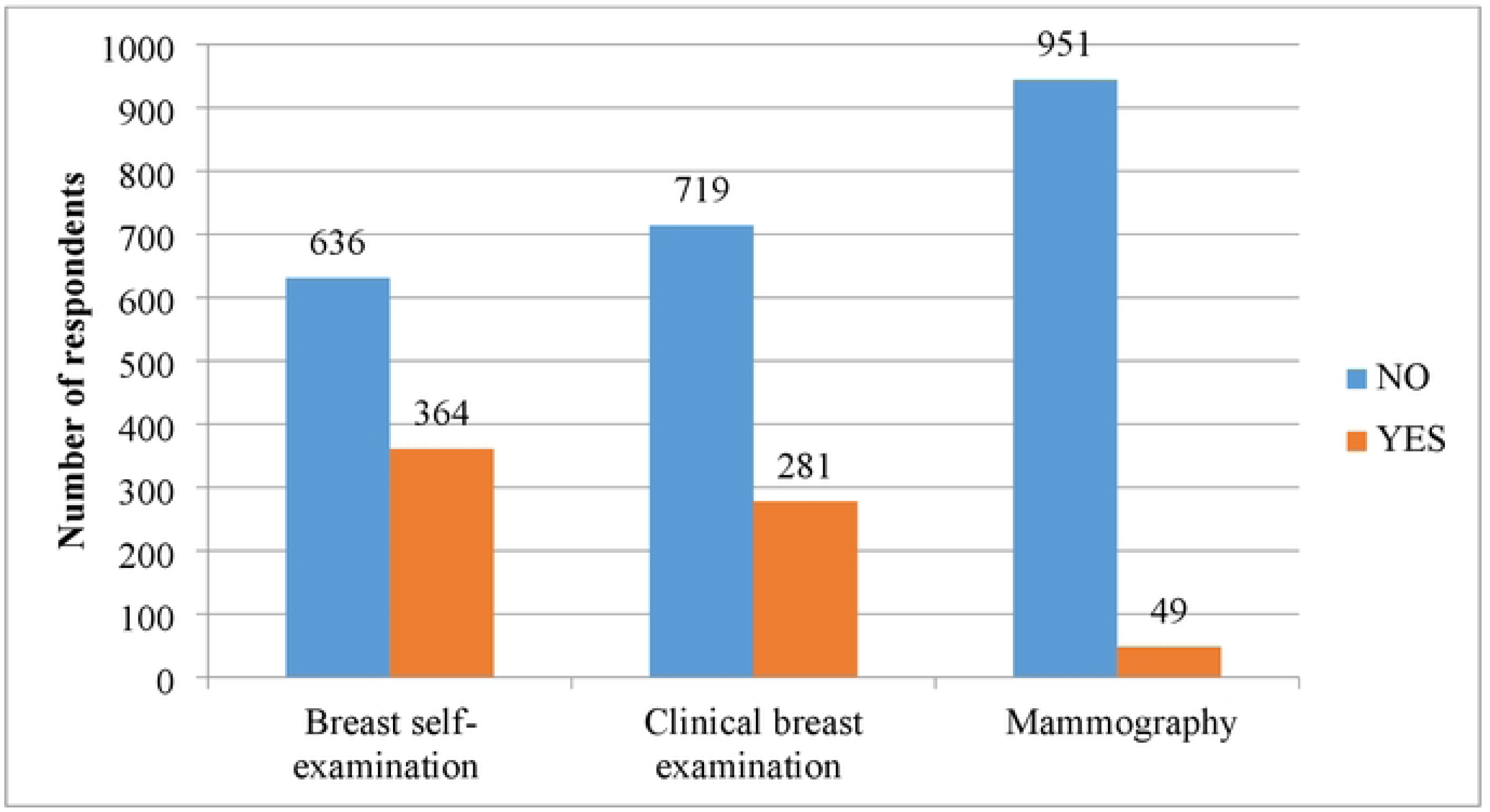
Types of breast examination among women aged `20` and above

### The practice of respondents related to breast self-examination

In this study, the prevalence of BSE was found to be 5.2%, with only 24(46.2%) participants reporting regular practice of BSE in accordance with the recommended guidelines. (Table-3)

**Table 3:** Practice of breast self-examination among women aged 20 and above in Sidama Region, South Ethiopia.

| Variable | Frequency | Percent |
| --- | --- | --- |
| **The practice of breast self-examination n=1000** |  |  |
| No | 948 | 94.8 |
| Yes | 52 | 5.2 |
| **Why do/did you perform BSE n=52** |  |  |
| Had previous breast problem | 5 | 9.6 |
| Fear of breast cancer from family | 5 | 9.6 |
| Recommended by a health professional | 20 | 38.5 |
| For early detection and treatment | 19 | 36.5 |
| Fear of developing breast cancer | 3 | 5.8 |
| **How often you practice BSE n=52** |  |  |
| Once in a week | 4 | 7.7 |
| Once in a month | 24 | 46.2 |
| Once in 3 month | 9 | 17.3 |
| Once in 6 month | 4 | 7.7 |
| When it comes to mind | 11 | 21.1 |
| **When do you perform BSE n=52** |  |  |
| Two to three days after menstruation session | 11 | 21.2 |
| When it comes to mind | 25 | 48.1 |
| A regular day of each month | 13 | 25 |
| Few days before menses | 1 | 1.9 |
| Any time during the month | 2 | 3.8 |

### Reason for not practice of breast self-examination

The majority of respondents, 948 (94.8%), had never practiced BSE. Among those who did not practice BSE, the primary reasons mentioned were the absence of breast problems, cited by 495(52.2 %) participants, and lack of knowledge on how to perform BSE, mentioned by 338(35.7%) participants. These factors were as the primary reasons for the low prevalence of BSE practice among the women in the study. (Figure-3)

**Figure 3:**
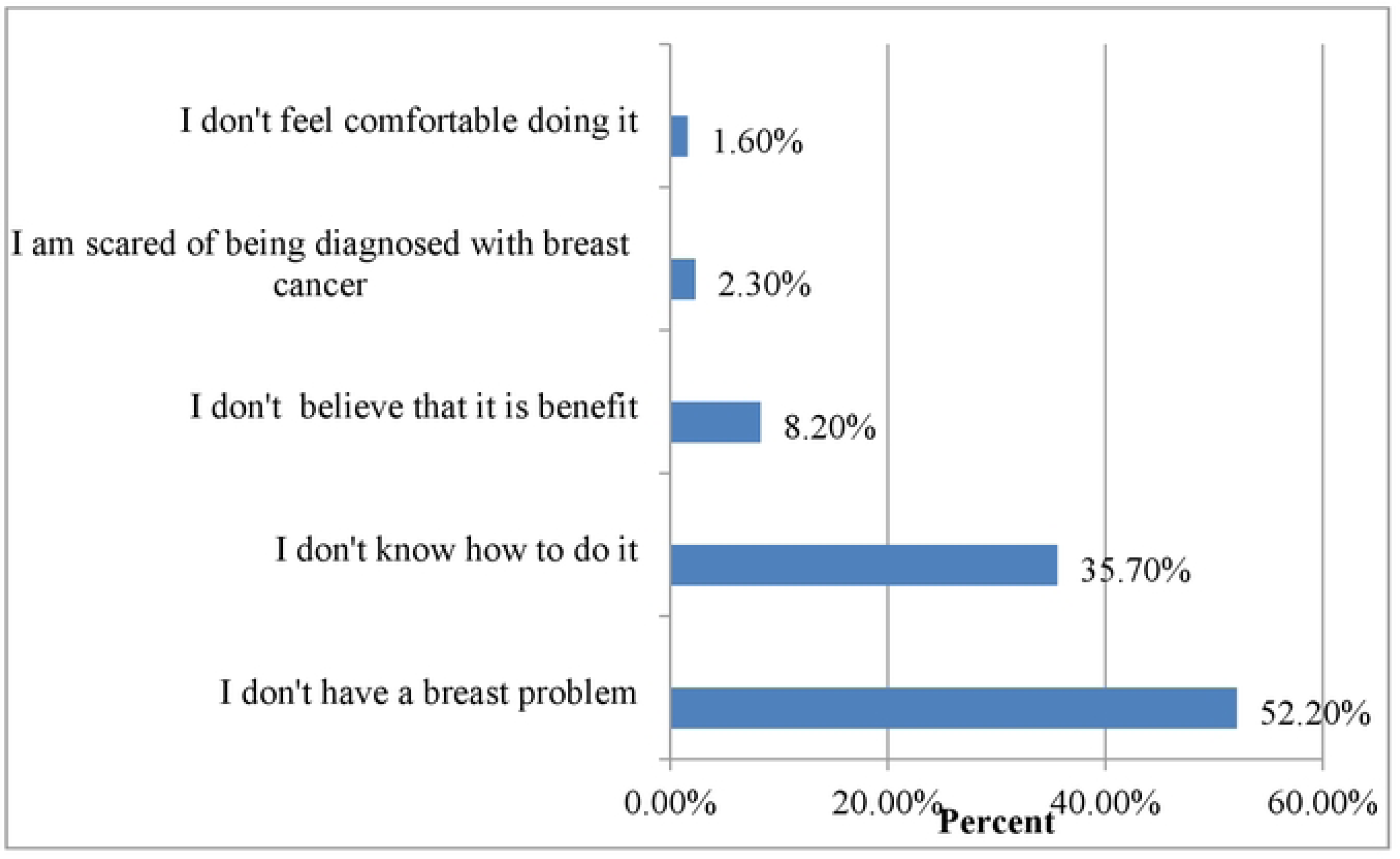
Reasons for not practicing BSE among women aged 20 and above in Sidama Region, South Ethiopia.

### Factors associated with breast self-examination practice

After adjusting for other factors, it was found that women’s knowledge about BSE for breast cancer, as well as three constructs of the health belief model (perceived susceptibility, perceived barrier, and perceived self-efficacy), were significantly associated with the practice of BSE.

Knowledge was identified as a significant predictor of BSE practice. Women with good knowledge were 4.5 times more likely (AOR=4.5, 95% CI: 1.4, 14.7) to practice BSE compared to women with poor knowledge. High perceived susceptibility was also associated with an increased likelihood of BSE practice, with women having a 2.7 times higher likelihood (AOR=2.7, 95% CI: 1.4, 5.2) compared to those with low perceived susceptibility.

On the other hand, high perceived barriers were associated with a decreased likelihood of BSE practice. Women with high perceived barriers were 70% less likely (AOR=0.30, 95% CI: 0.11, 0.8) to practice BSE compared to those with low perceived barriers. Additionally, women with high perceived self-efficacy were 3.9 times more likely (AOR=3.9, 95% CI: 2.0, 7.7) to practice BSE compared to women with low perceived self-efficacy (Table-4).

**Table 4.** Bivariable and multivariable logistic regression for factors associated with breast self-examination practice in Sidama Region, South Ethiopia.

| Variable | Practice of BSE |  | Odds ratio and 95% CI |  |
| --- | --- | --- | --- | --- |
|  | Yes (%) | No (%) | Crude | Adjusted |
| **Education status of women** |  |  |  |  |
| None formal education | 10(2.7) | 358(97.3) | 1 | 1 |
| Secondary education | 13(7.2) | 168(92.8) | 2.7(1.2,6.4)* | 1.3(0.43,3.9) |
| Tertiary and above | 9(17.3) | 43(82.7) | 7.5(2.8,19.5)** | 1.6(0.37,7.5) |
| **Educational status house head** |  |  |  |  |
| None formal education | 10(3.4) | 281(96.6) | 1 | 1 |
| Tertiary and above | 13(14.3) | 78(85.7) | 4.6(1.9,11.1)** | 1.1(0.28,4.13) |
| **Knowledge** |  |  |  |  |
| poor knowledge | 16(2.3) | 670(97.7) | 1 | 1 |
| Satisfactory knowledge | 29(10.1) | 258(89.9) | 4.7(2.5,8.8)** | 2.8(1.4,5.5)* |
| Good knowledge | 7(25.9) | 20(74.1) | 14.6(5.4,39.5)** | 4.5(1.4,14.7)* |
| **Perceived susceptibility** |  |  |  |  |
| Low perceived susceptibility | 19(2.7) | 694(97.3) | 1 | 1 |
| High Perceived susceptibility | 33(11.5) | 254(88.5) | 4.7(2.6,8.5)** | 2.7(1.4,5.2)** |
| **Perceived Benefit** |  |  |  |  |
| Low perceived benefit | 10(2.8) | 348(97.2) |  | 1 |
| High perceived benefit | 42(6.5) | 600(93.5) | 2.4(1.2,4.9)* | 0.8(0.39,1.9) |
| **Perceived barrier** |  |  |  |  |
| Low perceived barrier | 47(6.7) | 650(93.3) | 1 | 1 |
| High perceived barrier | 5(1.7) | 298(98.3) | 0.23(0.1,0.6)* | 0.30(0.11,0.8)* |
| **Perceived confidence** |  |  |  |  |
| Low perceived self-efficacy | 18(2.4) | 746(97.6) | 1 | 1 |
| High perceived self-efficacy | 34(14.4) | 202(85.6) | 6.9(3.8,12.6)** | 3.9(2.0,7.7)** |

\* Significant with P-value < 0.05, \*\*Significant with P-value < 0.001

## 6. Discussion

In this study, a Small proportion of respondents (5.2%) reported ever practicing BSE. This finding is similar to a study conducted in Adwa, in northern Ethiopia, which reported a BSE practice of being (6.5%) among participants (17). However, the BSE practice in our study was lower to other studies conducted in different areas. For instance, studies in Kaffa zone, southern Ethiopia (11%), west Gojam Zone, northwest Ethiopia (37.3%), Market women in southwest Nigeria (29.2%), Imo state Nigeria (51.9%), and rural area in Haryana, India (8.7%) showed higher BSE practice (10,11,21,22,23). This disparity could be attributed to the lack of community based awareness creation and screening programs for BSE in the study area. Additionally, differences in socio-demographic characteristics, such as educational background and low knowledge regarding the benefits of BSE, might contribute to the lower practice observed in our study.

Among those who had ever practiced BSE, only 24(46.2%) reported regular BSE practice. This finding contrasts with a study conducted in Kaffa Zone, where 73.07% of participants reported regular BSE practice (11). This difference might be due to variations in the number of study participants and socio-demographic characteristics, with female teachers potentially having better awareness of the benefits of regular BSE compared to the broader community.

The majority of respondents (94.8%) in our study had ever practiced BSE. The primary reasons mentioned were absence of breast problem (52.2%) and a lack of knowledge on how to perform BSE (35.7%). These results align with the study conducted in West Gojjam, where 53.2% of the participants cited the absence of breast problems and 30.6% mentioned lack of knowledge as reasons for not practicing BSE(10). This pattern could be attributed to cultural similarities in the study population, where healthcare seeking behaviors tend to prioritize treatment after illness rather than preventive activities (24).

In this study, 36.4% of the participants mentioned BSE as a primary screening method to detect breast cancer, followed by clinical examination at 28.1% and mammography at 4.9%. A study conducted among female health professionals in West Ethiopia reported higher values, with 64.3% being aware of BSE as a screening method, while 45.7% and 32.7% were knowledgeable about clinical breast examination and mammography, respectively (25). The difference in findings could be attributed to the nature of the population. The second study focused specifically on health professionals, who may possess better information and knowledge about breast cancer and BSE compared to the general population.

In this study, it was found that 56.5% of the respondents believed that early detection of breast cancer improves survival. However, a higher percentage of 85.7% was reported in a study conducted in Harmaya (26). The inconsistency in findings can be explained by the difference in study participants. The participants in the aforementioned study were health science students who are expected to have a higher level of knowledge regarding the breast cancer examination and its benefits compared to the participants in this study.

In our study, knowledge of participants emerged as a significant predictor of BSE practice. Participants with good knowledge were five times more likely to practice BSE compared to those with poor knowledge. These finding is consistent with a study conducted among household head women in western, and northern Ethiopia (25,27). Having Knowledge about breast cancer and BSE plays a crucial role in empowering women, boosting their confidence, and motivating them to perform BSE practices.

In the current study, women who had a high perceived susceptibility of breast cancer were three times more likely to practice BSE compared to those with low perceived susceptibility. This finding aligns with a different study conducted in a different area of Ethiopia(11,17).

Individuals with a high perceived susceptibility tend to personalize the risk of breast cancer, which increases their perceived threat and concerns. This heightened awareness and concern serve as motivators for them to perform BSE practice for early detection, ultimately aiming to improve the cancer outcomes of cancer management.

In this study, women who had a high perceived self-efficacy to perform BSE practice were four times more likely to practice BSE compared to their counterparts. This finding was consistent with studies conducted in North Ethiopia and Malaysia (17,28). Women with high perceived self-efficacy to perform BSE have a better understanding of the benefits of BSE, highly developed health-seeking behavior, and hold the belief that early diagnosis reduces the risk of breast cancer. Additionally, they are able to overcome perceived barriers to taking action. On the other hand, this finding suggested that a lack of skill in the performing of BSE is associated with limited or no BSE activity. In this study, a high perceived barrier to practicing BSE practice for breast cancer was associated with a 70% decrease in the likelihood of performing BSE compared to their counterparts. Similar findings were observed in studies conducted among Saudi and Turkish women (13,29). Women who have a low perceived barrier may possess a better knowledge on the benefits of BSE and hold strong beliefs that early detection can improve the outcome of breast cancer. This knowledge and belief system likely facilitate their engagement in BSE practices, as they are less hindered by perceived barriers.

### Limitations of the study

One potential concern of this study may be a recall bias of the respondents, which refers to the difficulty faced by women in accurately recalling past events when responding to questions related to knowledge and likert-scale assessements used to assess awareness and percpetions may have limitations in recalling specific details or accurately reflecting their experiences, which may be impact the reliability of their responses

## Conclusion

This study highlighted a significant low BSE practice compared to previous studies done in Ethiopia, with only a small number of women performing regular BSE. The participants demonstrated a lack of knowledge regarding BSE and breast cancer compared to findings from previous studies. The primary reason for not practicing BSE was lack of knowledge on how to properly examine their breasts. The study identified that women’s knowledge of BSE, as well as three constructs from the health belief model (perceived susceptibility, self-efficacy, and perceived barrier), were significant predictors of BSE practice. This finding underscores the importance of improving knowledge and addressing the barriers associated with BSE to encourage more women to engage in this self-screening practice.

The major reasons not to practice BSE. In this study we identified that knowledge of women with BSE for breast cancer and three health belief model constructs, that are perceived susceptibility, perceived self-efficacy, and perceived barrier, were significant predictors of BSE exercise.

## Acknowledgements

The authors hereby express their deep gratitude to Hawassa University, Vice President for Research and Technology Transfer Office, Research and Technology coordinator, office of College of Medicine and Health Science for allowing us to conduct this study. Sidama zone health department, Dale, Shebadino and Boricha woreda district Health offices, for their support and cooperation. We are also extremely grateful to the supervisor, data collectors, and women who participated in this study for their willingness to share their experiences of breast self-examination practices.

## Data Availability

The data used to support the findings of this study will be provided on request

## Authors’ Note

This study was approved by the Institutional Review Board of Hawassa University, College of Medicine and Health Sciences **(No. IRB/047/11, dated 26/02/2019)**. The participation of study participants was voluntary. Oral consent was obtained from all participants prior to their participation in the study

